# A systematic review of convalescent plasma treatment for COVID19

**DOI:** 10.1101/2020.06.05.20122820

**Authors:** Ville N. Pimenoff, Miriam Elfström, Joakim Dillner

## Abstract

**Background:** Transfusion of convalescent immune plasma (CP) is commonly used in epidemics. Several articles now describe clinical report data of CP for treatment of SARS-CoV-2-induced COVID-19 disease.

**Methods:** A systematic literature review was conducted using the NCBI curated COVID-19 related open-resource literature database *LitCovid* to identify studies using CP as treatment for COVID-19 patients. We retrieved and curated all COVID-19 related patient and treatment characteristics from previously reported studies. A Poisson model was developed to evaluate the association between age of the patients, older age being the most common risk factor for COVID-19 mortality, and recovery time since CP treatment using data extracted from the literature.

**Results:** From 18,293 identified COVID-19 related articles, we included ten studies reporting results of CP treatment for COVID-19 from a total of 61 patients. Decreased symptoms of severe COVID-19 and clearance of SARS-CoV-2 RNA were the most direct observations. We found that patients over the age of sixty who received CP treatment for COVID-19 had a significantly prolonged recovery estimated by viral clearance (from 10 to 29 days since first dose of CP) compared to younger patients, who recovered from the infection in less than a week after receiving CP treatment.

**Conclusions:** Limited published results on plasma transfusion treatment for COVID-19 disease with concomitant treatments suggest that CP therapy for COVID-19 is well tolerated and effective. First randomized clinical trial results, however, revealed no improvements in recovery time for elderly patients with severe COVID-19 between standard treatment alone and added with convalescent plasma. Accordingly, we argue that older patients may need a significantly longer time for recovery. Further randomized clinical trial data for COVID-19 with rigorous ethical standards is urgently needed.

## Background

Coronaviruses infecting humans and associated with severe atypical pneumonia (Severe acute respiratory syndrome coronaviruses 1 [SARS-CoV-1] and 2 [SARS-CoV-2], and Middle East Respiratory Syndrome coronavirus [MERS-CoV]) have caused three epidemics in the 21^st^ century^1–3^. There were 8,096 confirmed infections and 774 deaths in the SARS-CoV-1 outbreak in 2002 and 2,494 infections and 858 deaths in the MERS CoV outbreak in 2012^4,5^. The SARS-CoV-2 outbreak started in 2019 in China and has, so far, resulted in more than 6.5 million infections and more than 380,000 deaths^6^.

Passive immunization using convalescent plasma (CP) containing neutralizing antibodies, obtained from individuals that have recovered from the infection, was used during the SARS-CoV-1 and MERSCoV outbreaks^7,8^, and has also been proposed as a treatment option for COVID-19^9,10^. CP therapy may be most effective when given early during coronavirus infection and may actually cause harm to the patient if the plasma used has low antibody titers and the infusion is given late in the disease progression^11^.

Here we systematically review all available studies using convalescent plasma transfusion to treat COVID-19 and to estimate the association between age and COVID-19 recovery time since plasma transfusion.

## Material and Methods

### Systematic literature retrieval

To identify clinical studies using convalescent plasma as a treatment for COVID-19, a systematic literature review was conducted (June 3, 2020). Peer-reviewed studies were identified using inclusion criteria: **i)** keywords “***convalescent plasma***” with “***COVID-19***” and **ii)** reporting CP treatment of patients suffering from COVID-19, and **iii)** including the demographics and clinical characteristics listed in Table 1 for systematic review. To search systematically we used the comprehensive NCBI curated COVID-19 related open-resource literature database *LitCovid*^12^. Additional non-peer-reviewed scientific preprints were identified using the same inclusion criteria with databases http://www.medRxiv.orgs and http://www.biorxiv.org.

**Table 1.**
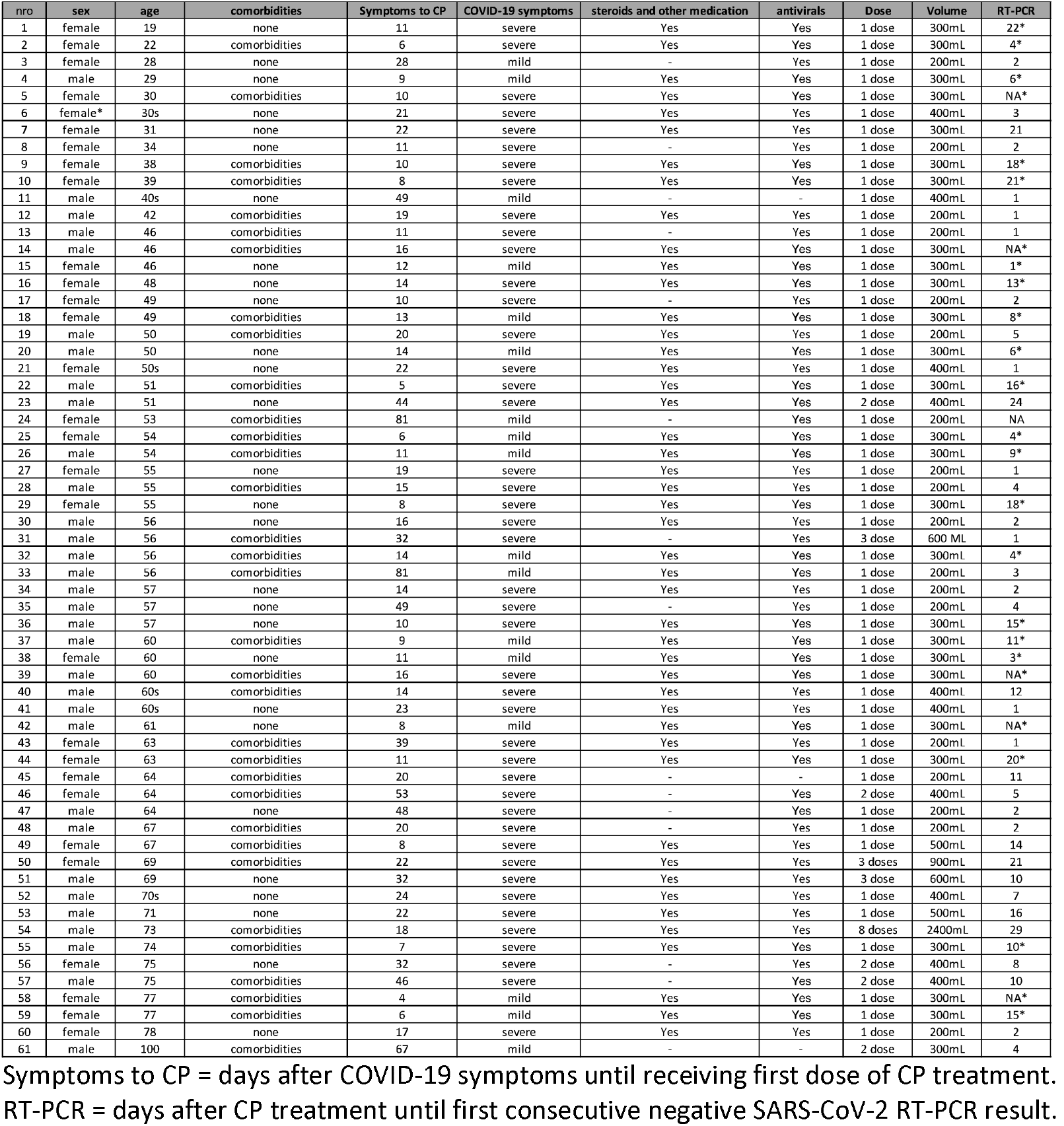
Clinical characteristics of the patients before and after convalescent plasma transfusion.

Initially, the literature search revealed 3,864 peer-reviewed articles on treatment strategies, therapeutic procedures, and vaccine development for SARS-CoV-2 infection and COVID-19 treatment (Figure 1). A total of 232 articles were identified that addressed the potential use of CP therapy for COVID-19, mostly by reviewing CP therapy results^13^ from previous infectious disease outbreaks such as influenza^14,15^, Ebola^16^ as well as SARS-CoV-1^17^ and MERS-CoV^8^. In addition, 203 non-peer-reviewed preprints mentioned CP therapy for COVID-19, five reported new clinical results and only one preprint had patient treatment data characteristics available.

**Figure 1.**
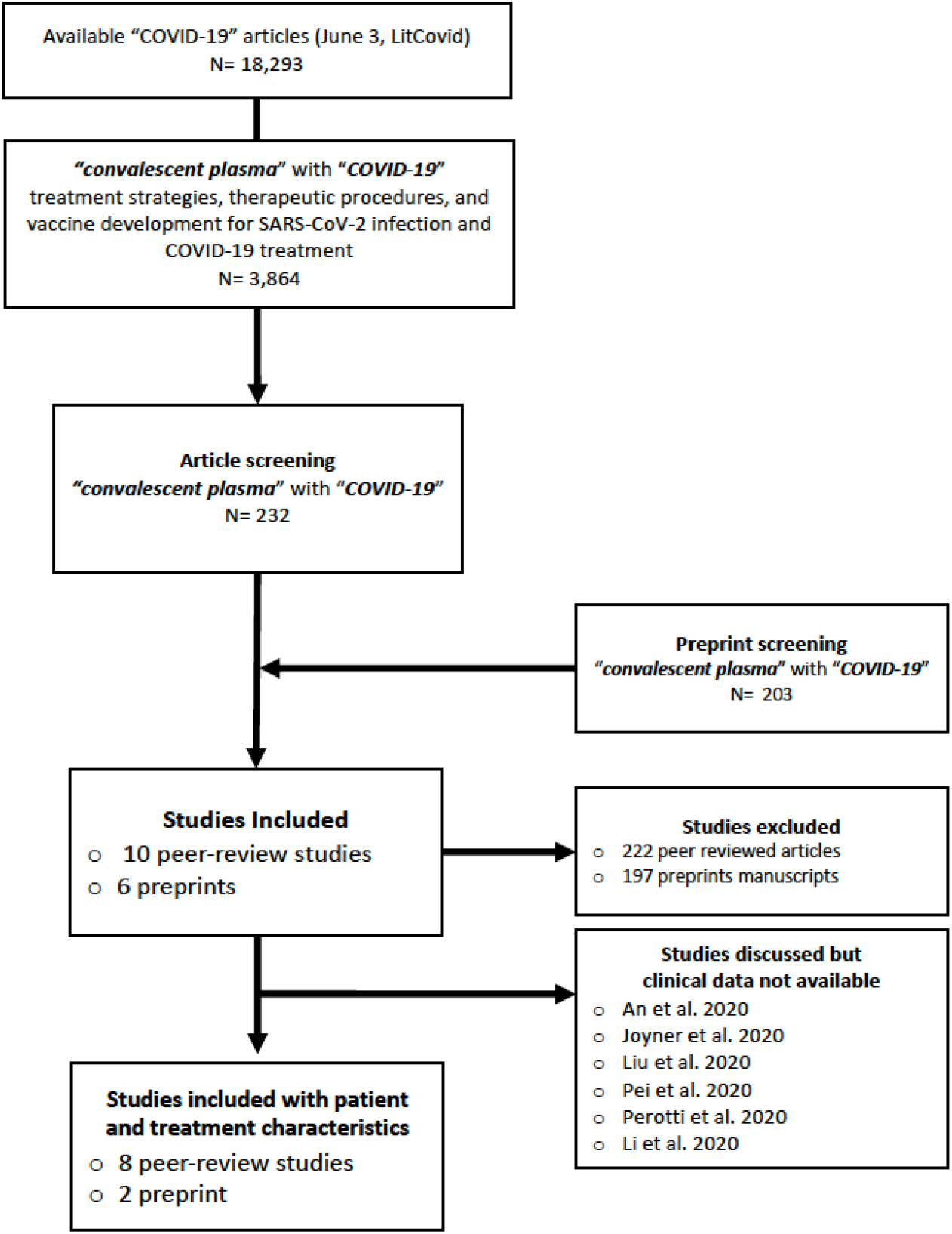
Flow chart of the systematic review study selection. Ten studies included in this systematic review are listed in Table 1, and studies discussed but without patient-level data available to be included in the analysis of this review are the following: An et al. 2020^47^; Joyner et al. 2020^35^; Liu et al. 2020^33^; Pei et al. 2020^36^; Perotti et al. 2020^46^ and Li et al. 2020^34^.

### Statistical analysis

In Table 1 we summarize the demographic and clinical characteristics retrieved from the studies and used in this review. The most important variables included in the analysis were gender, age, underlying health status, number of infusions and amount of plasma given, and recovery time after CP treatment, and the time since the onset of COVID-19 to receiving CP treatment for each patient. For the statistical model, we used demographic risk factor information for COVID-19 including age, gender, and underlying health status. For categorical age-related risk factor stratification a cut-off at 60 years was set following the definition that approximately 80% of COVID-19 related deaths have occurred among adults aged ≥60 years^18–20^. Previous reports also suggest that male gender and comorbidities such as hypertension and cardiovascular disease are risk factors for mortality for patients with COVID-19^18–21^, and thus were included in our model. Regarding CP treatment, the number of days between symptom onset and receipt of the first CP treatment was retrieved from all reported cases and included in the model. To estimate the safety of convalescent plasma for patients with COVID-19 all information about adverse events were retrieved from the studies included in this review. Finally, to estimate the efficacy of the CP intervention for COVID-19, the study endpoint was measured as the time to recovery in number of days since the CP treatment. Recovery was defined as the first consecutive negative result for SARS-CoV-2 RNA (i.e. CT value higher than 37) from mouth/nasal swab of the patient. If viral RNA results were not reported, we used the hospital discharge time point as a proxy for SARS-CoV-2 clearance. For the multivariate regression model, the time to recovery variable was stratified into two categories; i) 1–7 days and ii) more than 7 days since CP treatment. Notably, the use of other treatments such as steroidal and antiviral medication was also retrieved from the reviewed studies (Table 1). Prevalence ratios (PRs) were estimated with the described COVID-19 patient variables and using Poisson regression models with robust variance. All data analyses were carried out using R version 3.6.3 and related statistical packages.

## Results

### Literature review of the immune plasma antibody treatment

To evaluate the effect of immune plasma antibody treatment for COVID-19, our literature search identified ten studies (Table 1), which met our systematic literature review criteria, and reported results for CP therapy treatment for patients with confirmed COVID-19. Altogether these studies consisted of 61 patients from China, Korea and United States mostly with severe COVID-19 and reported detailed demographic and clinical characteristics of each patient receiving CP treatment^22–31^.

### Patients treated with immune convalescent plasma

Our systematic review included 61 patients between 19 and 100 years of age (29 women and 32 men), and who had experienced COVID-19 related symptoms for 1- 60 days before hospitalization (Table 1). Upon admission, clinical symptoms of COVID-19 such as ARDS (acute respiratory distress syndrome) and fever were observed in 72.1% (44/61) of the cases while the remaining 27.9% (17/61) of the patients we reported only with mild COVID-19 symptoms. All COVID-19 patients reviewed here were reported to have been confirmed by SARS-CoV-2 positive RT-PCR from nasopharyngeal/mouth swabs samples. Based on the available clinical data, most of the patients received antiviral medication such as arbidol, lopinavir-ritonavir and interferon alpha-1b, and more than 75% (46/61) of the patients received also other concomitant treatments such as steroids (e.g. methylprednisolone). Patients were selected to receive CP transfusion between 3 and 64 days after admission due to ongoing COVID-19 including respiratory difficulties or failure requiring mechanical ventilation despite antiviral and steroidal treatment, except in 27.9% (17/61) of cases with mild COVID-19 symptoms (Table 1). That is, patients received CP treatment between 4 and 81 days after the onset of COVID-19 related symptoms (Table 1). Nearly half of the patients, twenty-eight cases (45.9%), had no underlying medical conditions while twenty-five patients (41.0%) were at least hypertensive or diabetic, and the remaining eight patients (13.1%) had at least asthma, hypothyroidism, hyperlipidemia, bronchitis, Sjögrens syndrome, cancer of the esophagus or suffered from other pulmonary or cardiovascular disease. Transfusion of SARS-CoV-2 neutralizing antibody containing immune plasma was administered as a one-day treatment (i.e. one dose in Table 1) for 52 patients with a total of between 200mL and 500mL of CP for each patient. However, for nine patients, additional dose(s) of CP were infused up to a total of 300mL (1 patient), 400mL (4 patient), 600mL (2 patient), 900mL (1 patient) and even up to 2400mL (1 patient) of CP within a week, except one case, after the first dose due to rapid progression of pneumonic disease and no observed treatment response to the first dose of CP transfusion (Table 1). The plasma for recipient patients was obtained from the selected donors mostly within few days before the CP transfusion, if reported, and compatibility was controlled by evaluating the antibody titers and ABO blood type. Convalescent plasma donors had recovered from COVID-19 and were invited to donate plasma at least 10 days after recovery, with written informed consent. Donors had been diagnosed with COVID-19 and had a SARS-CoV-2 infection confirmed by RT-PCR from nasopharyngeal specimens. Donors tested negative for SARS-CoV-2 RNA after recovery and prior to plasma donation. Other respiratory infections, Hepatitis B, hepatitis C, human immunodeficiency virus (HIV) and syphilis were reported to have been tested following standard blood banking practices at the time of blood donation.

To investigate the effect of the CP intervention for COVID-19, we estimated that 50.8% (31/61) of the patients recovered within the first week and in 39.3% (24/61) of the cases patients recovered between 8 and 29 days after CP transfusion (Table 2A, see also Table 1). For 9.8% (6/61) of the patients the recovery data was not available (Table 1). Restricting the analysis to patients with RT-PCR results showed that the prevalence of viral RNA became negative in 66.7% (24/36) of the patients within the first week and in 30.6% (11/36) of the patient between 8 and 29 days after CP transfusion (Table 2B, see also Table 1). To further assess the differences in time to recovery after CP therapy for COVID-19 patients, we retrieved available patient and COVID-19 related treatment characteristics from all the studies reviewed here including gender, age, underlying health status and CP treatment time points since the onset of COVID-19 symptoms for each patient (Table 1). Our analysis revealed that patients over the age of sixty with COVID-19 who received CP treatment had a significant, nearly two-fold increased prevalence ratio of prolonged recovery of more than one week compared to younger COVID-19 patients who received CP therapy (Table 2A). These results were sustained and significant also when retaining only the COVID-19 patients with RT-PCR results (Table 2B) or exclusively among severe COVID-19 patients with RT-PCR results available (Table 2C).

**Table 2A.** Prevalence ratios between elderly risk group compared to younger COVID-19 patients (N = 61) and stratified by recovery time since convalescent plasma transfusion.

|  | < 60 years | ≥60 years | PRs | 95% CI |  |
| --- | --- | --- | --- | --- | --- |
| 1-7 days | 22 | 11 | ref | - |  |
| over 7 days | 9 | 13 | **1.92** | 1.02 - 3.58 | p= 0.042* |

Recovery stratified by median into between 1-7 or over 7 days (i.e. 8-29) after CP treatment until negative SARS-CoV-2 RNA detection.
Age < 60 includes patients with age ranging 19 to 57 years old.
Age > 60 includes patients with age ranging 60 to 100 years old.
Recovery time was not known for 6 patients with COVID-19 and were excluded from the analysis (see Table 1).
\*Wald test p value adjusted by sex, comorbidity and time of CP administration since the onset of COVID-19 symptoms.

**Table 2B.** Prevalence ratios between elderly risk group compared to younger COVID-19 patients (N = 36) and stratified by recovery time since convalescent plasma transfusion.

|  | < 60 years | ≥60 years | PRs | 95% CI |  |
| --- | --- | --- | --- | --- | --- |
| 1-7 days | 16 | 2 | ref | - |  |
| over 7 days | 8 | 9 | **2.22** | 1.21 - 4.06 | p= 0.010* |

Recovery in 1-7 or over after CP treatment until negative SARS-CoV-2 RNA detection.
Age < 60 includes patients with age ranging 28 to 57 years old.
Age ≥60 includes patients with age ranging 60 to 100 years old.
Salazar et al. study (N=25, <sup>30</sup>) lacking recovery time estimation by SARS-CoV-2 RNA and one additional patients with no recovery time reported <sup>27</sup> were excluded from the analysis.
\*Wald test p value adjusted by sex, comorbidity and time of CP administration since the onset of COVID-19 symptoms.

**Table 2C.** Prevalence ratios between elderly risk group compared to younger patients exclusively with severe COVID-19 (N = 31) and stratified by recovery time since convalescent plasma transfusion.

|  | < 60 years | ≥60 years | PRs | 95% CI |  |
| --- | --- | --- | --- | --- | --- |
| 1-7 days | 13 | 2 | ref | - |  |
| over 7 days | 7 | 9 | **1.97** | 1.05 - 3.69 | p= 0.035* |

Recovery in 1-7 day or over after CP treatment until negative SARS-CoV-2 RNA detection.
Patients with mild COVID-19 symptoms and Salazar et al. study (N=25, <sup>30</sup>) lacking recovery time estimation by SARS-CoV-2 RNA were excluded from the analysis.
Age < 60 includes patients with age ranging 30 to 57 years old.
Age > 60 includes patients with age ranging 60 to 78 years old.
\* Wald test p value adjusted by sex, comorbidity and time of CP administration since the onset of COVID-19 symptoms.

No significant differences were observed in recovery time after CP treatment between gender (Table 3A), between patients with and without underlying chronic diseases (Table 3B) nor between patients receiving CP therapy within the first three weeks and after three weeks since the onset of COVID-19 symptoms (Table 3C). These results were consistently sustained also when analyzing only the COVID-19 patients with RT-PCR results (results not shown). The reviewed studies mostly used clinical symptoms and CT scanning to monitor the initial response and recovery of the patient from severe pneumonia in the first weeks after CP therapy. Across the studies, antiviral treatment was mostly given until the SARS-CoV-2 RNA was not detected in the nasopharyngeal/throat swab sample. Patients allergic to plasma content, positive for HBV, HCV or HIV or with uncontrolled bacterial infection were reported to have been excluded in most of the identified studies.

**Table 3A.** Prevalence ratios between gender of COVID-19 patients (N = 61) and stratified by recovery time since convalescent plasma transfusion.

|  | women | men | PRs | 95% CI |  |
| --- | --- | --- | --- | --- | --- |
| 1-7 days | 13 | 18 | ref | - |  |
| over 7 days | 13 | 11 | **0.78** | 0.45 - 1.37 | p= 0.388* |

Recovery in 1-7 day or over after CP treatment until negative SARS-CoV-2 RNA detection.
Recovery time was not known for 6 patients with COVID-19 and were excluded from the analysis.
\*Wald test p value adjusted by age, comorbidity and time of CP administration since the onset of COVID-19 symptoms.

**Table 3B.** Prevalence ratios between non-comorbid and comorbid COVID-19 patients (N = 61) stratified by recovery time since convalescent plasma transfusion.

|  | No<br>comorbidities | comorbid | PRs | 95% CI |  |
| --- | --- | --- | --- | --- | --- |
| 1-7 days | 18 | 13 | ref | - |  |
| over 7 days | 9 | 15 | **1.27** | 0.73 - 2.21 | p= 0.388* |

Recovery in 1-7 day or over after CP treatment until negative SARS-CoV-2 RNA detection.
Recovery time was not known for 6 patients with COVID-19 and were excluded from the analysis.
\*Wald test p value adjusted by sex, age and time of CP administration since the onset of COVID-19 symptoms.

**Table 3C.** Prevalence ratios between early and late administration of CP among COVID-19 patients (N = 61) and stratified by recovery time since convalescent plasma transfusion.

|  | early CP | late CP | PRs | 95% CI |  |
| --- | --- | --- | --- | --- | --- |
| 1-7 days | 19 | 12 | ref | - |  |
| Over 7 days | 17 | 7 | **0.73** | 0.35 - 1.56 | p= 0.423* |

Recovery in 1-7 day or over after CP treatment until negative SARS-CoV-2 RNA detection.
Recovery time was not known for 6 patients with COVID-19 and were excluded from the analysis.
\*Wald test p value adjusted by sex, age and comorbidity of the patients.

## Discussion

The main finding of this review is that CP treatment was consistently safe and created an immediate recovery response in about half of the patients reviewed. This signal of shorter time to recovery was significantly associated to patients under the age of sixty. No severe adverse effects of the CP therapy were observed, though, concomitant treatments were also reported in all ten studies included. That is, all patients recovered, 51% (31/61) within the first week after CP treatment. Notably, however, for nine patients, additional doses of CP up to 300–900mL, and even up to 2400mL, were needed, but a full recovery was observed in these cases as well within days after the last dose of CP transfusion.

Accumulating evidence shows a strong age-related difference in disease development for COVID-19 patients, with an estimated 80% of COVID-19 related deaths among adults aged ≥60 years^19,20^, and longer time to recovery for older patients is therefore expected. An intriguing observation however is that despite the recently established higher mortality rates for COVID-19 among men and individuals with comorbidities, particularly hypertension^19–21^, these risk characteristics were not found in this review to be related to recovery time after CP therapy. The first sign of recovery reported in these studies was that the clinical symptoms of COVID-19 such as fever, cough, shortness of breath and chest pain largely improved or disappeared within days after CP transfusion. Another clinical observation was that patients also showed different degrees of pulmonary recovery in CT scans in the days and weeks after CP transfusion, although reporting of clinical improvements was heterogeneous across the ten studies.

The most robust measure to estimate the CP treatment response of the COVID-19 patients across the ten studies was deemed to be the RT-PCR of SARS-CoV-2 RNA from nasopharyngeal and throat swab samples. Even so, for a number of cases the viral RNA results were not available^30^, and hence, for those we used the reported patients hospital discharge time as a proxy for expected clearance of SARS-CoV-2 RNA. We estimated that 51% (31/61) of the patients recovered within the first week and all 55 patients with recovery data available within 29 days after CP treatment. When reported, the neutralizing antibody titers of the patients was sustained or increased in the first week immediately after CP transfusion. Conforming, it is argued that the level of neutralizing antibodies in the plasma is critical for the effectiveness of the treatment^32^. Interestingly, however, a recent study speculates that CP treatment may actually require an incubation of more than a week after transfusion before manifesting survival effect for severe COVID-19^33^.

Altogether, the results reviewed here suggest that CP therapy for COVID-19 is safe and effective, as all patients were alive and virtually all discharged, however the safety and effectiveness of convalescent plasma for patient with COVID-19 is difficult to fully evaluate based on the very limited and heterogeneous case report data available. Particularly, all COVID-19 patients reviewed here received in parallel antiviral and other medications, and thus, these concomitant treatments prevented direct estimations of the likely benefits or harms of CP treatment. Nevertheless, no adverse effects were observed in any of the ten convalescent plasma therapy COVID-19 studies included in this review. In agreement, first randomized clinical trial study published among patients with severe COVID-19 reported only two patients out of 52 with adverse event likely related to CP transfusion, and both cases recovered in hours with standards treatment^34^. This study also reported that convalescent plasma transfusion added to standard treatment for severe COVID-19 patients and compared with standard treatment alone did not significantly improve the recovery from the disease in four weeks of follow-up^34^. Another ongoing study of 5000 COVID-19 patients treated with convalescent plasma also emphasize that the treatment is safe, as thus far, they report only two patients with severe adverse effects certainly related to the CP transfusion^35^. Pei and colleagues^36^, on the other hand, have reported one COVID-19 patient with a severe anaphylactic shock after receiving convalescent plasma. Most recently, an interventional single arm study from Italy reported reduced mortality for COVID-19 patients treated with convalescent plasma^37^. However, patient and clinical data from these four studies is not fully available, and thus, could not be included in the analyses of this review. Taken together, there is clearly a lack of randomized clinical trial results of CP treatment for COVID-19.

Importantly, however, previous experience with CP therapy for coronavirus-related severe pneumonic disease, such as SARS-CoV-1 and MERS-CoV, already pointed a note of caution for the treatment and argued that CP therapy may be most effective when given soon after onset of the disease^8^. In agreement, an improved intervention outcome was observed among SARS patients who received CP within the first two weeks of disease onset^38^. Contrary to this, we did not find any association between recovery time and time for receiving CP therapy either at early or at late time point since the onset of COVID-19 (Table 3C). Nevertheless, recent reports estimating the feasibility and risks of the convalescent plasma for larger scale treatment of pandemic COVID-19 note that current ongoing trials of CP for COVID-19 may underestimate the risks of the treatment if administered at low antibody titer at late time point of developing severe COVID-19 disease^11,39^. Indeed, there is still very limited data available to estimate the effects of CP plasma treatment for COVID-19 patients at large scale^34^. Numerous clinical trials have already been initiated and are currently ongoing worldwide (at least 48 reviewed in ^40^) and both FDA and the European Commission have recently established COVID-19 trial guidelines^32,41,42^. An important concern, however, has been recently addressed that COVID-19 clinical trials may be currently implemented largely due to urgent treatment needs but at the expense of rigorous scientific and ethical standards^43,44^. One immediate risk being an imperfect evaluation of the convalescent plasma safety and compatibility, which can lead into an increased risk of adverse events or transmission of infectious disease agent. Accordingly, It has been recently emphasized that the optimal time for collecting CP should be determined by the time and level of total antibody production in convalescent patients^45^. Another important caveat related to plasma transfusion treatment for COVID-19 patients, which warrants future evaluation, is the possible transfusion-associated circulatory overload, a common serious adverse effect of transfusion^11^. Furthermore, it is suggested that there may be a risk for complement-mediated tissue damage in convalescent plasma treatment^11^. Lastly, the risk for antibody-dependent disease enhancement during CP treatment for COVID-19 requires further evaluation using randomized clinical trial data^11,39^. Taken together, current reports on convalescent plasma for treating COVID-19 is limited by lack of representation of patients in the early phase of infection, as well as confounding from multiple overlapping therapies and small patient numbers^34^. Randomized clinical trial data is needed for comprehensive evaluation of CP treatment safety and feasibility, however, not treating patients with severe COVID-19 for clinical trial purposes may need a rigorous ethical evaluation.

### Limitations

Despite the significance of our results, our study has a number of limitations. There is only limited data available, thus far, for a comprehensive analysis of the safety and feasibility of CP therapy for COVID-19. A massive number of large scale clinical trials are ongoing, but preliminary data is still not available^33–36,46^. Ideally, all the patients would have been followed with the same criteria for the COVID-19 onset and recovery. However, we have curated all eligible studies and used measures available across all the studies included in the review. Moreover, we re-analyzed our data after excluding the study lacking RT-PCR results for SARS-CoV-2 RNA post-transfusion^30^. The results presented in this review remain significant also in these sensitivity analysis.

## Conclusion

Most available intervention results of CP transfusion with high titer SARS-CoV-2 antibody to treat COVID-19 disease is limited but suggest that the therapy is safe and effective for the treatment of patients suffering from COVID-19 disease. In contrast, however, a recent randomized clinical trial comparing treatments among elderly risk-group patients with severe COVID-19 did not observe significant difference in time of recovery between standard treatment alone and added with convalescent plasma. Accordingly, we argue that older age may be an underlying characteristic of COVID-19 patients associated with prolonged recovery time after CP transfusion. Moreover, patient’s comorbidities, gender or time of CP transfusion since the onset of COVID-19 symptoms seems to have limited effects on recovery time after CP treatment.

## Data Availability

All data used in this article is publicly available.

## Acknowledgements

We thank Johan Ursing and Maria Matl for helpful discussions. This research was financially supported by an unrestricted donation from Creades. The funder did not have any role in the interpretation of the results, nor in the preparation of this manuscript.

